# Income-based differences in health care utilization in relation to mortality: Trends in the Swedish population between 2004-2017

**DOI:** 10.1101/2023.03.31.23287996

**Authors:** Pär Flodin, Peter Allebeck, Ester Gubi, Bo Burström, Emilie E. Agardh

## Abstract

**Background:** Despite universal health care, socioeconomic differences in health care utilization (HCU) persist in modern welfare states. The aim of this study is to assess income-based differences in utilization of primary- and specialized care in relation to mortality for the Swedish general population (>15 years old) between 2004 and 2017.

**Methods and Findings:** Using a repeated cross-sectional register-based study design, data on utilization of i) primary-ii) specialized outpatient- and iii) inpatient care, as well as iv) cause of death, were linked to family income and sociodemographic control variables. HCU and mortality for all-disease as well as for the five disease groups causing most deaths were compared for the lowest (Q1) and highest (Q5) income quintile using logistic regression. We also analysed income-related differences in the number of health care encounters ≤5 years prior to death.

In 2017, for all diseases combined, Q1 utilized marginally more primary- and specialized outpatient care than Q5 (adjusted odds ratio [OR] =1.07, 95% CI: 1.07, 1.08; OR 1.04, 95% CI: 1.04, 1.05, respectively), and considerably more inpatient care (OR=1.44, 95% CI: 1.43, 1.45). The largest relative inequality was observed for mortality (OR 1.78, 95% CI: 1.74, 1.82). This pattern was broadly reproduced for each of the five diseases.

Time trends in HCU inequality varied by level of care. Each year, Q1 (vs. Q5) used more inpatient care and suffered increasing mortality rates. However, usage of primary- and specialized outpatient care increased more among Q5 than in Q1. Finally, Q1 and Q5 had similar number of encounters in primary- and inpatient care ≤5 years prior to death, but Q1 had significantly fewer outpatient encounters.

**Conclusions:** Income-related differences in the utilization of primary and specialized outpatient care were considerably smaller than for mortality, and this discrepancy widened with time. Facilitating motivated use of primary- and outpatient care among low-income groups could help mitigate the growing health inequalities.

## Introduction

Low socioeconomic status (SES) is associated with higher mortality rates, and more years lived with disability and disease (1). Despite universal health care, socioeconomic differences in health care utilization (HCU) persist in modern welfare states (2). Reducing avoidable socioeconomic inequalities is a high priority in public health, and provision of health care in proportion to need is one cardinal strategy. In Sweden, as in many countries, the law ordains that “…those who have the greatest need for care must be given priority” (3). However, the degree to which this has been accomplished is not conclusively determined.

In a recent systematic review of 57 studies from high income countries, Lueckmann et al. (2) report that in the majority of studies, higher SES was associated with higher utilization of specialized outpatient care for a given self-reported health status. The findings of primary care utilization were more heterogeneous, and the authors concluded that inequities were more common in specialized outpatient care compared to primary care. Hence, Lueckmann et al. (2) pointed out that “.. a distinction of medical appointments between primary and specialized [outpatient] care is necessary when analysing socioeconomic inequalities in physician visits, because the results differed greatly according to the type of doctor and the type of service.”

To evaluate the effects of public policy and health care reforms on inequality in HCU and mortality, analysis of time trends are informative (4). Although Sweden was successful in reducing inequalities e.g., by providing universal health care and free education, relative inequalities in mortality have increased since 1990 (5). The Swedish health care system has during the same period undergone major changes, such as the marketization of primary care (spurred by the Primary Health Care Choice Reform in 2010) and a recent increase in virtualization of primary care. Although previous research indicates that marketisation increased inequalities in HCU (6) the evidence is sparse and incomplete (7). Similarly, studies of trends in inequalities in utilization of specialized care show mixed results (8,9), and updated, large scale studies are lacking. Importantly, to what extent trends in socioeconomic difference in HCU relates to mortality inequalities is not well studied.

The Scandinavian health registers provide unique opportunities to investigate this. In Sweden, national population health care registers are regularly updated (10), and microdata on HCU and death can be linked to social databases through civic registration numbers. This enables analysis of socioeconomic differences and utilization of primary- and specialized outpatient and inpatient care as well as mortality for the general population over time.

The aim of this study is to assess time trends of income-related differences in use of different levels of health care in relation to mortality between the years 2004 and 2017. To do this we compared the lowest (Q1) with the highest income quintile (Q5) with regard to mortality and the utilization of primary care, specialized outpatient care, and inpatient care, respectively. We studied each of the five disease groups that account for the largest number of deaths in Sweden 2017 (cardiovascular disease, neoplasms, neurological disease, chronic respiratory disease, and diabetes), as well as all-disease. Specifically, we addressed the following questions: 1) How do low- and high-income groups differ with regard to HCU and mortality in 2017 and over time? 2) To what extent do income-related differences in HCU and mortality vary across disease groups? 3) How do the low- and high-income groups differ in numbers of health care encounters ≤5 years prior to death?

## Methods

### Study population and data sources

The study population comprised all individuals 16 years and older living in Sweden any year between 2014 and 2017, that were registered in the Longitudinal Integrated Database for Health Insurance and Labour market Studies (11) by Statistics Sweden (Table 1). Data on specialized out- and inpatient HCU (from the National Patient Register, NPR) and mortality (from the Cause of Death Register, CDR) were obtained from the Swedish National Board of Health and Welfare (NBHW). Since primary care data is not available at the national level, we contacted the 21 Swedish counties maintaining electronic records of diagnoses in primary care. Seventeen counties provided time series covering one or more years (S1 Table). Whereas the national health registers provided by NBHW have well documented and high population coverage (10), primary care data lacked estimates of coverage rates, which likely vary by region and over time. Hence, crude rates of primary care utilization could partly be affected by increasing coverage rates and should be interpreted with caution.

**Table 1.** Study population. By type of register (national health registers from NBHW, or primary care data), time period, and income group (total population, Q1, and Q5).

| Type of register | Year(s) | Total population:<br>N | Total population:<br>Age (std) | Q1: N | Q1: Age (std) | Q5: N | Q5: Age (std ) |
| --- | --- | --- | --- | --- | --- | --- | --- |
| **NBHW**<br>(specialized care and cause of death) | **All years** | 105 343 853 | 48.49 (19.41) | 21075466 | 48.49 (19.41) | 21063689 | 48.49 (19.41) |
|  | **2017** | 8001909 | 48.74 (19.62) | 1600742 | 48.74 (19.63) | 1600073 | 48.74 (19.62) |
|  | **2011** | 7625859 | 48.12 (19.62) | 1525750 | 48.12 (19.62) | 1524841 | 48.12 (19.62) |
|  | **2004** | 7086141 | 48.49 (19.05) | 1417981 | 48.49 (19.05) | 1416834 | 48.49 (19.05) |
| **Primary care** | **All years** | 77812350 | 48.16 (19.36) | 15598476 | 47.87 (19.20) | 16678951 | 48.20 (19.33) |
|  | **2017** | 7142796 | 48.63 (19.59) | 1436285 | 48.64 (19.55) | 1456728 | 48.57 (19.56) |
|  | **2011** | 5841894 | 47.82 (19.53) | 1178215 | 47.61 (19.41) | 1239690 | 47.85 (19.50) |
|  | **2004** | 4101156 | 47.83 (18.91) | 803496 | 46.94 (18.51) | 962576 | 48.02 (18.92) |

For disease-specific analyses, we selected the five disease categories causing the highest number of deaths in Sweden in 2017, which together were responsible for 84% of all deaths (12). The disease groups were (with percentage of the total number of deaths in parenthesis): cardiovascular disease (38%), neoplasm (28%), neurological disease (8%), chronic respiratory disease (5%) and diabetes and chronic kidney disease (4%). Diseases were classified using tenth version of the WHO International Classification of Diseases (ICD-10) codes in accordance with the disease groups defined by the Global Burden of Disease project (S2 Table). Whereas NBHW define specialized care and underlying cause of death by ICD-10, diagnoses in primary care were classified using ICD-10-P which we mapped to ICD-10. Since ICD-10-P is less detailed than the ICD-10, certain disease groups in the primary care contain marginally broader disease categories than the GBD target groups.

Data on HCU and mortality were individually linked to register based data on family income, sex, birth year, residence county, country of origin, and civil status, as provided by Statistics Sweden.

### Income Measures

The study population was grouped by income ranks based on equivalised disposable family income. Family income referred to wages, capital returns, self-employment, pensions, and social benefit, after taxes were deducted (11). For each studied year, an individual’s family income was averaged across the 1-5 preceding years, and the resulting variable was subsequently used for income ranking. Individuals with a family income of zero or below (<2% of the population) were excluded from analysis due to the heterogeneity of this group (13). SES groups were defined as quintiles of time averaged family income, stratified by sex and birth year. Thus, at a given year, each income quintile contained approximately the same number of individuals and were balanced with regard to sex and age compositions.

### Statistical analysis

Crude rates of HCU per 100 000 person / year were calculated for each disease group and for each of the three levels of care. Only main diagnoses were considered. Mortality rates were based on underlying cause of death. For main analyses, we used the number of unique individuals, where an individual was counted once per disease group, calendar year and type of data source (i.e., primary care, outpatient, inpatient or death). Given the varying coverage rate of primary care register data, the counties contributing to the population denominator varied across the years, why it ranged from 4.1 million individuals in 2004, to 7.1 million in 2017. The population denominator used for the national health register data ranged from 7.1 million individuals in 2004 to 8.0 million in 2017 (Table 1).

We conducted multiple logistic regressions to calculate adjusted odds ratios (OR) of HCU or death, comparing the lowest (Q1) and highest (Q5) income quintiles. The model adjusted for country of origin, civil status, sex, age and age squared.

To infer time trends in ORs, we regressed log transformed OR on time. To account for uncertainty of the OR, we conducted bootstrapping analysis by simulating new time series (1000 resamples with replacements) drawn from a normal distribution with mean and standard deviation equal to the log transformed ORs. For each simulated time series, we performed linear regressions (GLM), and inferred 95% CI from the sampling distribution of the estimated beta coefficients.

Finally, we investigated whether the income groups differed regarding the number of health care visits during a five-year period prior to death. More specifically, for each year and each disease group, we specified a linear regression model of health care encounters by level of care, using income quintiles as independent variables with Q5 as the reference. We controlled for age, sex, county, and country of origin.

Analyses were carried out in SAS 9.4 and in Python 3.6 using standard data science packages such as *statsmodels*. All code is available at https://github.com/parflo/TrendsInMortalityHCUInequality.git.

The study was approved by the Regional Ethical Review Board, Stockholm, Sweden (DNR: 2018/1339-31/5, 2018/2292-32, 2019-02185, 2021-00657 and 2022-03111-02).

## Results

For the analyses of mortality and utilization of inpatient and outpatient care, we investigated 105.3 million person-years at risk. For primary care, we had access to data on 77.8 million person-years at risk. Outcomes in the cohort amounted to 1.3 million deaths, 9.9 million year-unique individuals in inpatient care, 38.4 million in outpatient care, and 45.9 million in primary care (Table 1).

Fig 1 shows OR for utilization of primary care, outpatient care, inpatient care, and mortality for each of the disease categories in 2017. Considering all-disease, those with lowest income (Q1) utilized slightly more primary care (OR =1.07, 95% CI: 1.07, 1.08) and outpatient care (OR=1.04, 95% CI:1.04-1.05) compared to those with the highest income (Q5). However, Q1 utilized considerably more inpatient care (OR=1.44, 95% CI: 1.43, 1.45) and had even higher death rates (OR= 1.78, 95% CI: 1.74, 1.82) (Fig 1, S4 Table).

**Fig 1.**
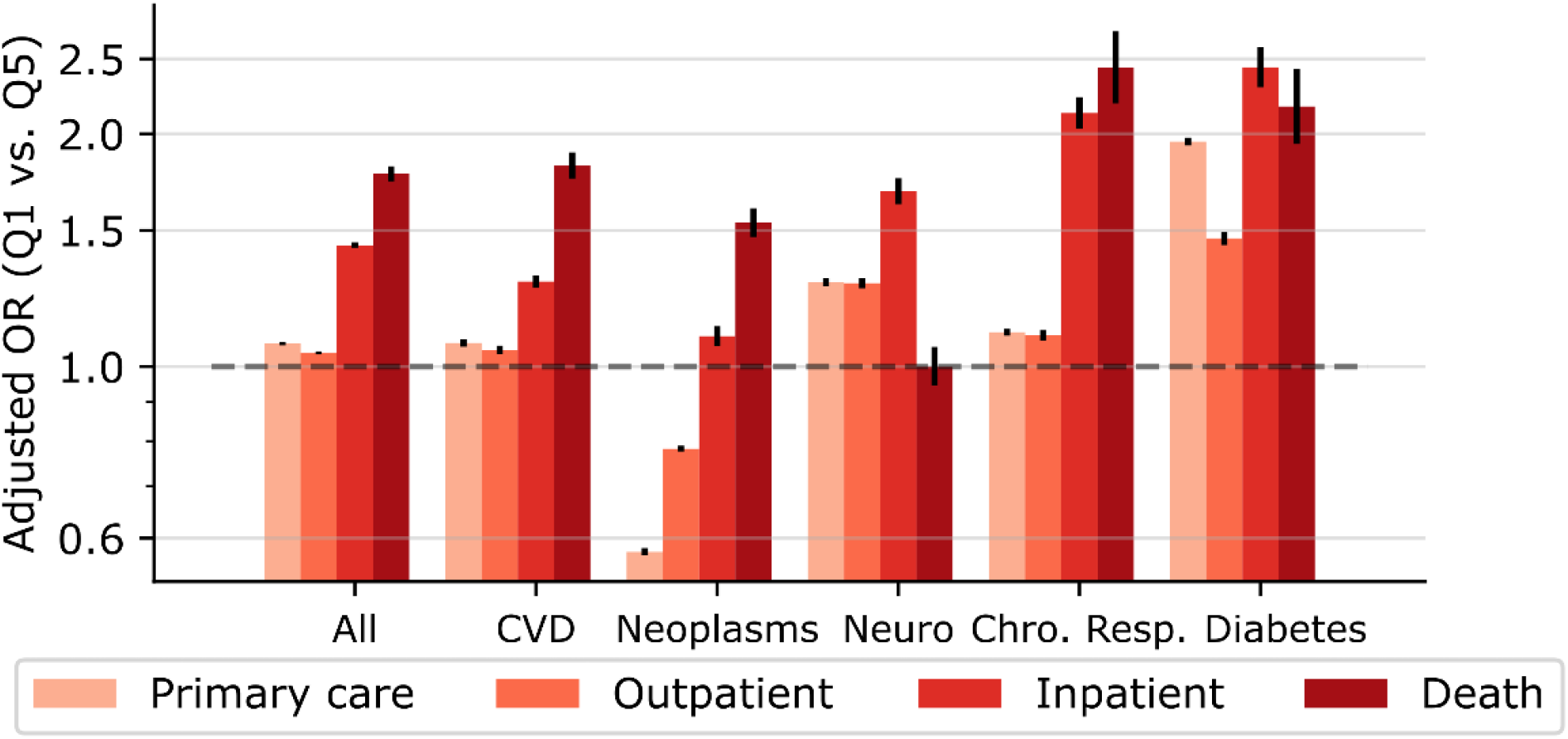
Adjusted odds ratios of HCU and mortality comparing the lowest with the highest income quintile, in year 2017. All: any ICD-10 code; CVD: cardiovascular disease; Neuro: neurological disease; Chro. Resp.: Chronic respiratory disease; Diabetes: Diabetes and kidney disease.

Similar patterns of income-related HCU and mortality were observed for each investigated disease. For both cardiovascular disease and neoplasm, ORs were lowest for utilization of primary and outpatient care and were considerably larger for inpatient care. The largest relative difference was found for mortality. Interestingly, use of primary and outpatient care due to neoplasms were lower in Q1 compared to Q5 (primary care: OR = 0.58, 95% CI 0.57-0.58; outpatient care: OR = 0.78, 95% CI 0.78-0.79) (Fig 1, S4 Table).

Fig 2 shows time series of rates of HCU and mortality for Q1, Q5, and for the total study population. For all the six disease categories, the yearly rates of unique individuals in primary and outpatient care are increasing. Rates of inpatient care plateaued around 2012 for most disease categories and decreased by the end of the study period. All-disease mortality rates for Q5 and the total population declined throughout the study period but stayed relatively unchanged for Q1. Cardiovascular disease contributed most to decreasing mortality rates in the total population (S3 Table).

**Fig 2.**
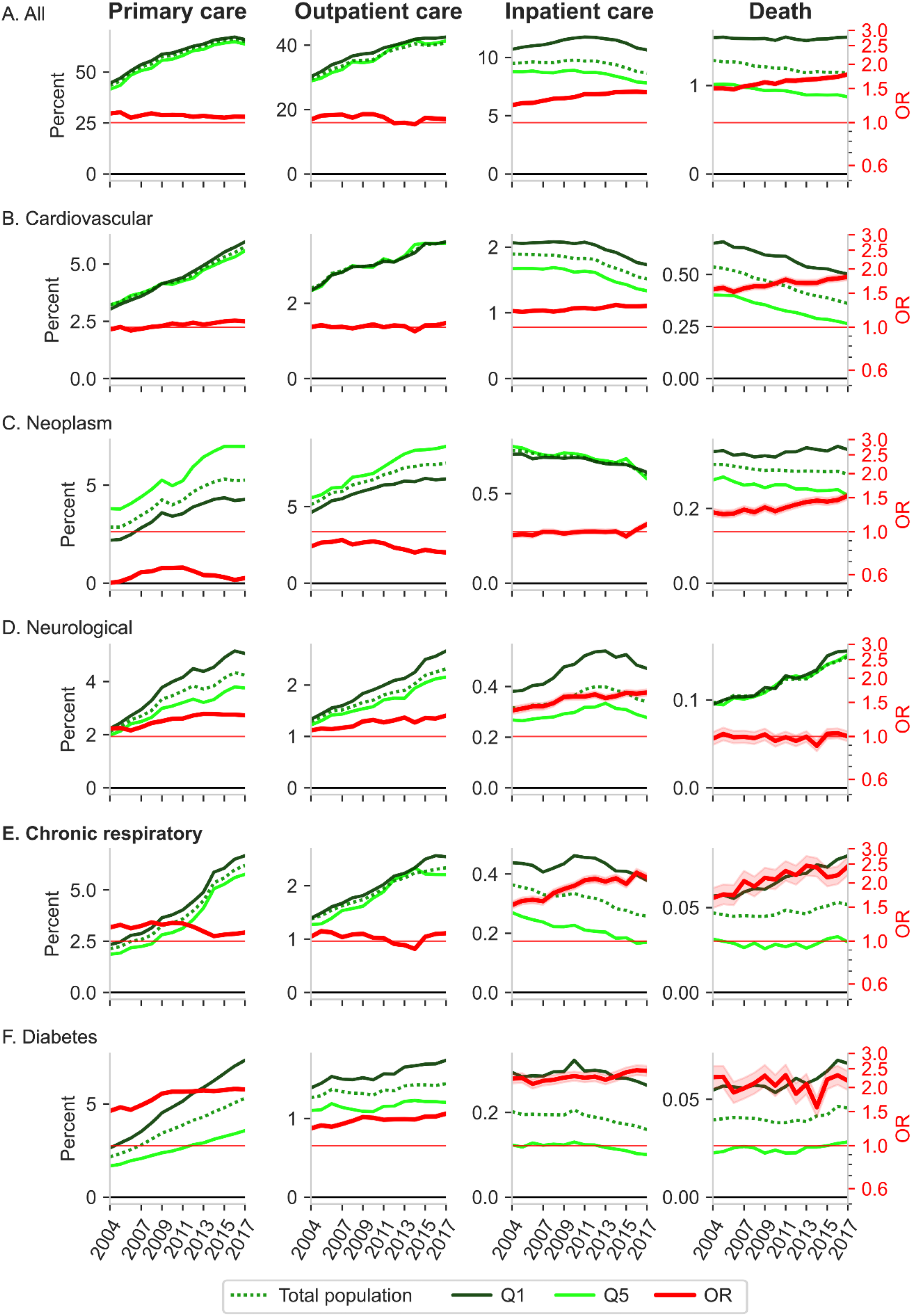
Time courses (2004-2017) of rates and adjusted odds ratios of HCU and mortality. HCU (yearly rates of unique individuals) and mortality rates, by income group and for the total study population (green colors). Rates are presented in percentages for display purposes. Adjusted ORs (95% CI) comparing Q1 with Q5 are in red. Numerical values of rates per 100 000 are found in S3 Table.

Fig 2 also shows time series of adjusted OR comparing Q1 with Q5. We observe an overall pattern of slightly decreasing ORs in primary- and outpatient care, but increasing ORs in inpatient care and mortality, for most disease categories.

Zooming in on the time-trends of OR, Fig 3 displays the beta estimates (i.e., the slope) of log transformed ORs regressed on time, which denotes the rate of change in the relative differences between Q1 and Q5. Time trends of ORs for all-disease followed a similar pattern as the one observed for 2017. Thus, for each year, Q1 (relative to Q5) displayed increasing rates of inpatient care, and even larger increase in mortality rates, but decreasing rates of primary- and especially outpatient care. For most of the specific disease categories, including cardiovascular disease and neoplasms, we observed a similar pattern of HCU and mortality trends (Fig 3, S4 Table).

**Fig 3.**
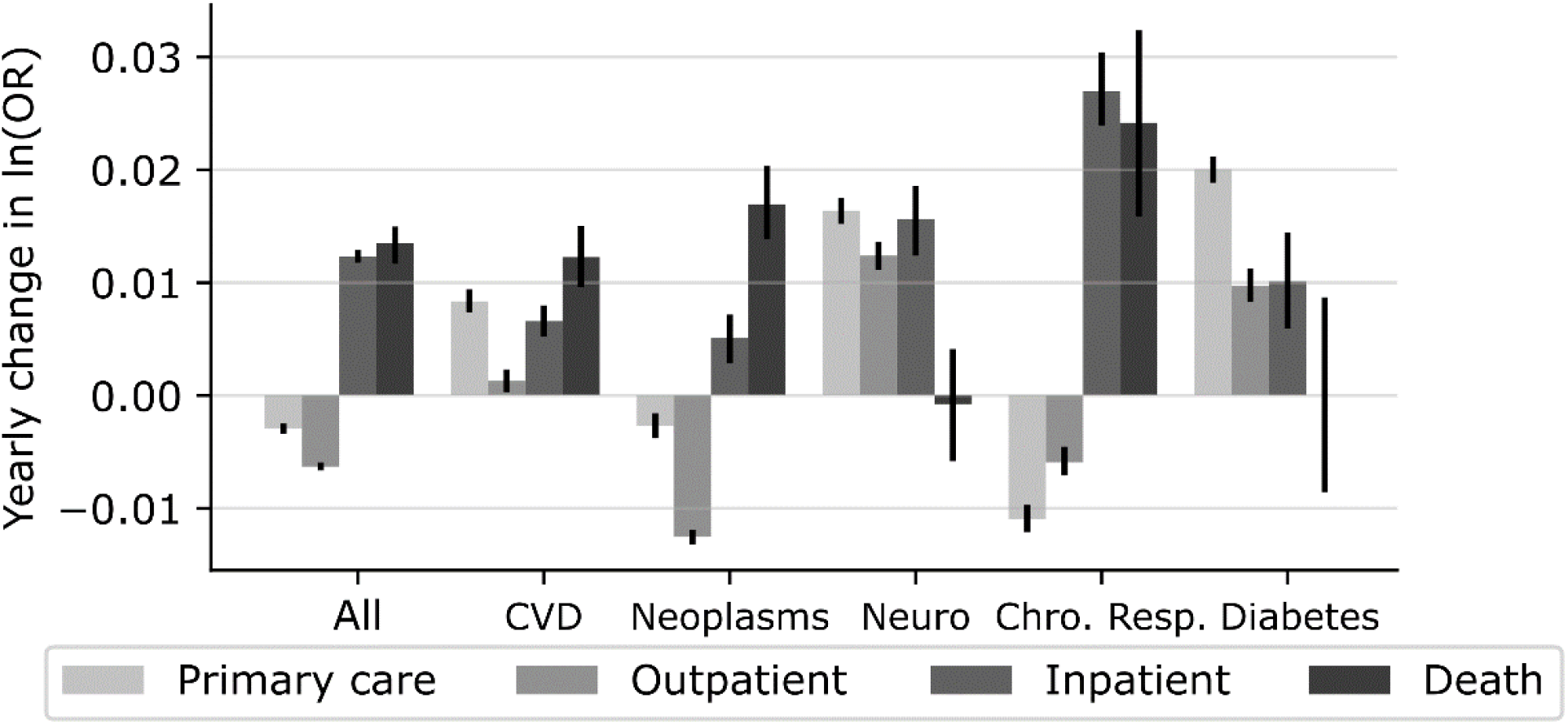
Yearly change in log OR. Bars show the beta-coefficients (i.e., rates of change or “slopes”) estimated by linear regression of the log transformed time-series of OR regressed on years (2004-2017). Error bars denotes 95% CI. For numerical values, see S4 Table.

Fig 4 shows income-related difference in number of disease specific health care encounters five years prior to death. In four of the five disease categories, Q5 had significantly more visits in outpatient care compared to Q1 and Q3 (Fig 4, S5 Table). For cardiovascular disease, Q1 had -23.9% (−32.1%, -15.7%) fewer outpatient encounters relative to Q5, and for neoplasm -13.5% (−19.1%, -7.8%) fewer. We also observed similar but smaller and more variable effects of income-related differences in number of primary care encounters. Estimates for respiratory disease and diabetes were associated with wider confidence intervals due to fewer deaths. Finally, income-related differences in inpatient care were small and largely non-significant for most of the disease groups most of the years.

**Fig 4.**
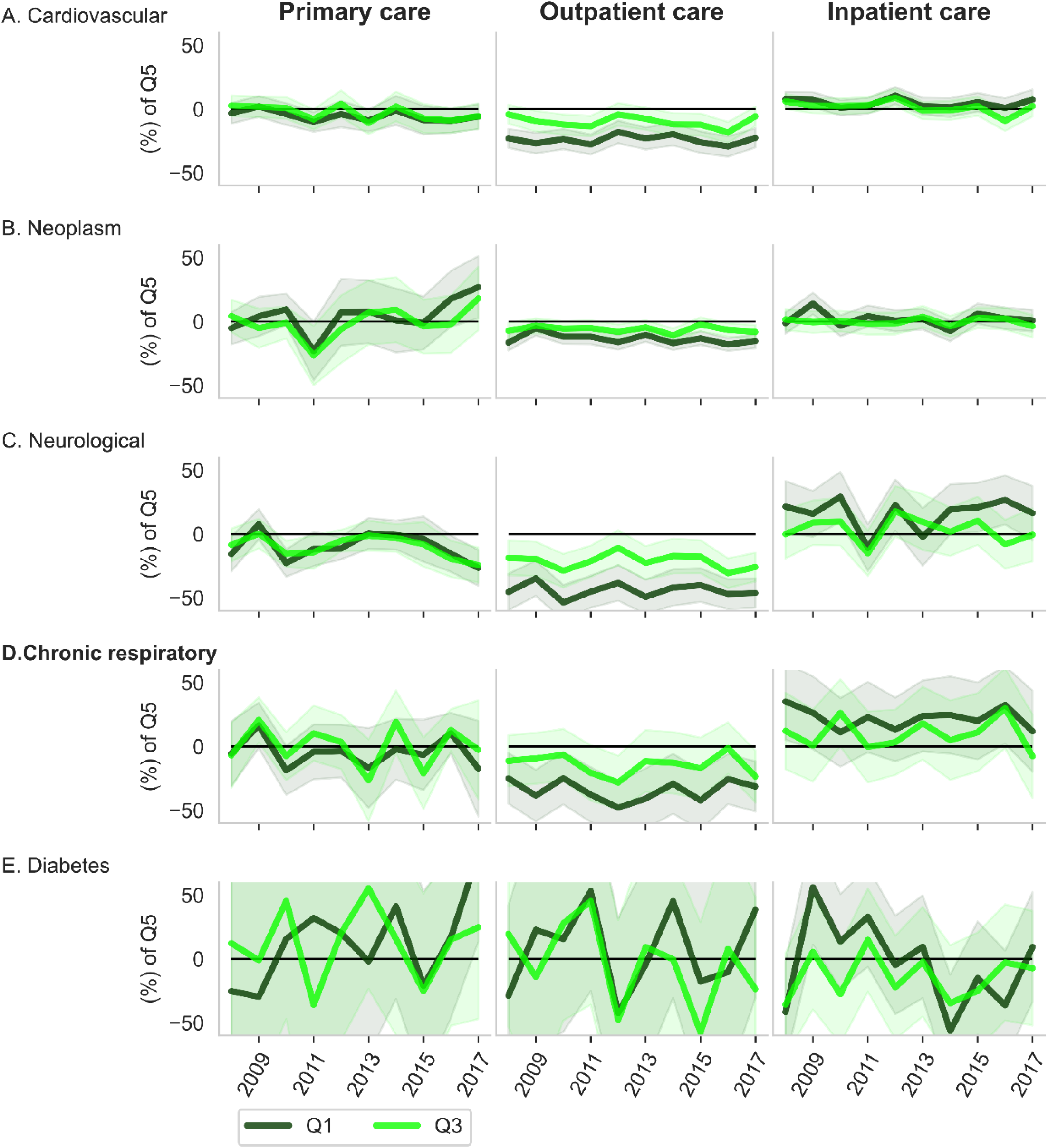
HCU before death. Income-related group differences in number of cause specific health care visits within 5 years prior to death, for death occurring during the period 2008-2017. Encounters of the lowest income quintile (Q1) and the middle-income quintile (Q3) are presented as percentages of the number of encounters by Q5.

## Discussion

In this study we analysed time trends of income stratified HCU and mortality. For all diseases combined, the more “severe” the outcome, the larger the inequalities. Compared to the highest income quintile, the lowest income quintile had 7 % higher odds of utilizing primary care, 4 % higher odds of utilizing specialized outpatient care, 44 % increased odds of hospitalization, and 78 % higher odds of dying, in year 2017. If HCU were in proportion to need, we would expect higher HCU among those in the lowest income quintile. In other words, the large inequality in mortality rates were not reflected in correspondingly large group differences in HCU, and the discrepancy between large inequality in mortality and lower in HCU was further widened each year.

### Trends in HCU inequalities

Our findings of income-based HCU inequalities are in line with previous survey studies. In Italy, Petrelli et al. found that high income earners utilized both more primary and specialized outpatient care, but less inpatient care, compared to low-income earners (14). This pattern is well documented in a wide range of high income countries (2,15,16), although there are a few older studies that only partly confirmed this (17,18).

Analogous to our findings, a longitudinal Norwegian survey study showed that socioeconomic differences in primary care utilization diminished with time between 1986 and 2006 (19). Moreover, high SES groups were more prone to utilize outpatient care, after adjustment of self-reported need. Finally, the income-based ratio of inpatient care was stable through the period, with considerably higher rates among low-income groups (19). Similarly, in a Swedish sample from year 1996/1997, Burström found an inverse income gradient related to having the need, but not seeking medical care (8).

It is noteworthy that the association between SES and HCU in part depends on the operationalization of HCU. Lueckmann et al. found that studies that defined HCU in terms of the probability of having visited a physician (“yes or no”), reported larger inequalities in favour of high SES groups, than studies reporting on frequencies (i.e., the number of health care encounters), or more rarely, conditional frequencies (meaning the number of encounters given the individual had visited a physician at least once). Hence Lueckmann et al. suggested that HCU inequalities would best be tackled by removing barriers in initial access to specialized outpatient care for deprived groups (2).

The importance of adjusting for need when analysing socioeconomic differences in HCU is widely acknowledged (20). “Need” may be conceptualized as the capacity to benefit from care. Need is commonly measured either as self-reported general health status, or as self-reported presence of disease. Among the studies listed by Lueckmann, the majority employed the former. However, for population wide register data, measures of subjective health are typically absent (as in [6]), which is also the case in our study. Instead, we compared income-related differences in HCU with group differences in mortality. Thus, the income-related HCU differences detected in the current study are likely due to a combination of differences in prevalence, access to care, and in care seeking behaviour independent of prevalence. The relative importance of each of these mechanisms is however unknown to us. This could hamper comparisons with previous findings of HCU inequalities.

Taken together, studies on HCU inequalities in high income countries mainly report higher HCU of specialized outpatient care among high SES groups, and higher rates of hospital admission among low SES groups. Results of primary care utilization have been more ambiguous, but our findings show that high income earners utilize primary care almost to the same extent as Q5, despite their substantially lower utilization of inpatient care and odds of dying. Although more updated analyses of trends are rare, the available evidence suggests that our findings of increasing use of outpatient care among high SES is not unique for Sweden.

### Trends in mortality inequalities

Overall, previous studies of income-based inequalities in mortality rates have reported trends comparable to our findings. Kondo et al. measured the income (Q1 vs Q5) ratio in mortality rates of the Swedish working age population between 1994 and 2004 and reported a yearly widening of mortality rates of 11% (5), and a more recent study report similarly (21). In Norway, Kinge et al. (22) observed an increasing income-related gap in life expectancy from 2005 to 2015 among individuals aged 60 to 69 years. Comparing four Scandinavian countries, Brønnum-Hansenfi concluded that the “Nordic countries displays the same overall pattern of increasing gap between income quartiles in mortality [between 1997 and 2017]” (23).

In this study, we observed that the largest inequalities in mortality were for chronic respiratory disease, followed by diabetes and cardiovascular disease. No income-related differences in neurological mortality rates were observed, in line with previous findings of larger health inequalities for more ‘‘preventable” diseases (1). If using the population attributable fractions (i.e., the percentage of disease burden that are explained by avoidable risk factors) provided by GBD (12) as a proxy for “preventability”, we observe a clear (albeit not perfect) relationship to inequality magnitudes: in Sweden in 2017, the population attributable fraction in mortality were 99.9% for diabetes, 81% for cardiovascular disease, 66% for chronic respiratory disease, 40% for neoplasm and 23% for neurological disorders.

Relative inequalities in mortality tend to increase as the overall mortality rates in the population declines (1). This is also what we found. Between 2004 -2017, all-disease mortality decreased from 1285 per 100 000 to 1143 per 100 000 in the general population. Although both Q1 and Q5 displayed decrease in all-disease mortality rates, the relative inequality increased from OR=1.50 to OR=1.74. However, increased relative inequalities are probably due to a combination of many factors, including social drift, socioeconomic differences in lifestyle, exposures to risk factors, and HCU.

### HCU and possible explanations for increased inequality in mortality

From a public health perspective, understanding the causal factors driving health inequalities is important since these provide potential targets for interventions. However, explaining the origin of the observed increase in mortality inequalities is challenging, and differences in HCU likely only play a partial role.

Income-related differences in utilization of primary- and outpatient care could be due to differences in both prevalence and care seeking behaviour, and here we cannot confidently disentangle the two. In contrast, income-related differences in inpatient care are likely driven almost exclusively by differences in prevalence of the conditions requiring hospital admission, a claim that is further substantiated by the observed corresponding magnitudes of mortality differences. It is tempting to speculate that given perfect horizontal equity (i.e., equal treatment for equal need), any relative group differences in utilization rates of primary- and outpatient care should be of similar magnitude as differences in inpatient use and mortality. However, the fact that we found substantially larger OR for mortality and inpatient care could possibly be due to many reasons. For instance, if low SES individuals would be more prone to more severe conditions (e.g., comorbidities) for which primary- and outpatient care are not a primary point of entry, one would expect larger relative group differences in inpatient care and death. To better determine this, future studies should use proper control for group differences in medical need.

### Health care availability

Although financial barriers to access to care have been largely removed in modern welfare states, the so-called *inverse care law* suggests that “availability of good medical care tends to vary inversely with the need of the population served” ([1] p 459, Hart 1971). An increasing number of private health providers and the free right to establish care centres in any location has in recent years reduced the number of child and maternal care units in poorer areas where the need is greater (24). In addition, availability is further skewed in favour of high SES HCU due to the increase in private insurances and out of pocket health care spending. However, since our study is based on records of publicly financed health care usage, privately paid care is omitted from our analyses. If anything, including it would likely further accentuate the observed discrepancy between income-based differences in HCU and mortality.

### The fundamental cause theory

The *fundamental cause theory* states that “SES embodies an array of resources, such as money, knowledge, prestige, power, and beneficial social connections that protect health no matter what mechanisms are relevant at any given time” (25 p28). HCU is one important mechanism by which people can protect and promote their health. It is well documented that high SES is associated with higher adherence to a range of preventive health programs, e.g. cervical cancer screenings (26) and vaccinations (27). Taken together, the current study adds to the growing body of evidence of an inverse care law (28), where the healthy and wealthy utilize primary and special care more than the low-income earners, adjusting for differences in need (either inferred from self-reports, or as in this study, -indicated by mortality).

### Strengths and limitations

To our knowledge, this is the first study to present time trajectories of inequalities both in mortality rates and health care utilization, covering the Swedish adult population for more than a decade. This was possible due to a unique, comprehensive database of linked microdata on socio demographics, cause of death, and health care utilization in specialized care, as well as on primary care in a majority of the Swedish counties.

Most previous studies on SES differences in HCU rely on survey data. Although these provide valuable information on self-reported need of care, survey studies are typically based on just a small fraction of the general population, interviewed at only a few time points. National population registers, on the other hand, provide (near) full population coverage, and enables unbiased definitions of income groups. Finally, the continuously recorded HCU register data enabled detection of time trends with high precision.

This study presents a bird’s-eye view of inequalities in HCU and morality over time at the expense of a more fine-grained description of patterns of group differences in HCU, which would be valuable when attempting to intervene the mechanisms causing avoidable health inequalities. For instance, pooling relatively heterogeneous diseases into broad disease categories masks the more extreme and possibly preventable income-related differences in HCU. Further, our metrics of HCU (i.e., yearly health care prevalence) tells little about inequalities in the treatments provided.

Another limitation is the unknown and time dependent coverage of primary care data. Coverage is likely higher in the more population dense counties, which typically also coincide with higher income gradients and higher concentrations of high-income individuals. If true, this could partly explain the relatively high primary care utilization among high income earners. However, similar patterns of high HCU among high income earners are also observed in the outpatient care, for which data coverage has been consistently high (29).

### Future studies

Further studies should investigate socioeconomic differences in HCU patterns with higher resolution in terms of disease categories, treatments, and patient flows (30). Our study indicates that a promising target for reducing health(care) inequalities is to facilitate access to primary- and outpatient care for low-income groups. Increased knowledge on how to promote care seeking related to neoplasms, cardiovascular disease, and chronic respiratory disease in low SES groups would be particularly valuable, considering their high prevalence and the large discrepancies between income-related differences in HCU and income-related differences in mortality for these disease categories.

## Conclusion

In this study we found major discrepancies between income-related differences in health care utilization and income-related differences in mortality, which widened with time. While high income earners utilized an increasing share of primary and outpatient care, inequality in mortality increased each year. This was especially noticeable for neoplasms. The relative underutilization of primary and outpatient care among low-income earners should be addressed to mitigate the trend of increasing health inequalities.

## Data Availability

Data cannot be shared publicly because of personal integrity. Data are available from the Swedish National Board of Health and Wellfare and Statistics Sweden, for researchers who meet the criteria for access to confidential data.

## Abbreviations

*HCU*: Health care utilization
*SES*: Socioeconomic status
*Q1*: Lowest income quintile
*Q5*: Highest income quintile

## Acknowledgments

We are grateful to all providers of register data, and in particularly to the providers of regional primary care data. We also thank Hugo Sjöqvist, Lode Van Der Velde and Dorien Beeres for valuable discussions on the data analyses, and Kasra Zarei for language proofreading.

## Supporting information

**S1 Table.** The year from which primary care data is available from each county.

| County | Inclusive start year |
| --- | --- |
| 01 | 2004 |
| 03 | 2010 |
| 05 | 1999 |
| 06 | 2012 |
| 08 | 2011 |
| 09 | 2013 |
| 10 | 2010 |
| 12 | 2004 |
| 14 | 2000 |
| 17 | 2014 |
| 18 | 2002 |
| 19 | 2017 |
| 20 | 2005 |
| 21 | 2010 |
| 22 | 2011 |
| 23 | 2010 |
| 25 | 2012 |

**S2 Table.** ICD definitions of disease groups.

| *Disease group* | *ICD-10 codes* |
| --- | --- |
| All-disease | * (all ICD codes) |
| Cardovascular disease | B33.2-B33.24, D86.85, G45-G46.8, I01-I01.9, I02.0, I05-I09.9, I11-I11.2, I11.9, I20-I21.6, I21.9-I27.0, I27.2-I28.9, I30-I38.0, I39-I41.8, I42-I43.8, I44-I44.8, I45-I52.8, I60-I64, I64.1, I65-I83.93, I86-I89.0, I89.9, I95.0-I95.1, I98, I98.8-I99.9, K75.1, R00-R01.2, Z01.3-Z01.31, Z03.4-Z03.5, Z13.6, Z52.7, Z82.3-Z82.49, Z86.7-Z86.79, Z94.1-Z94.3, Z95-Z95.9 |
| Neoplasms | C00-C07, C08-C19.0, C20, C21-C21.8, C22-C22.4, C22.7-C23, C24-C26.1, C26.8-C26.9, C30-C30.1, C31-C33, C34-C34.92, C37-C37.0, C38-C39.9, C40-C41.4, C41.8-C41.9, C43-C45.2, C45.7, C45.9, C47-C4A, C50-C50.629, C50.8-C52, C53-C54.3, C54.8-C56.2, C56.9-C58.0, C60-C64.2, C64.9-C69.92, C70-C70.1, C70.9-C73, C74-C75.5, C75.8-C79.9, C80-C81.49, C81.7-C81.79, C81.9-C85.29, C85.7-C86.6, C88-C90.32, C91-C93.7, C93.9-C95.2, C95.7-C97.9, D00-D24.9, D26.0-D39.9, D4-D49.9, E34.0, K51.4-K51.419, K62.0-K62.3, K63.5, N60-N60.99, N84.0-N84.1, N87-N87.9, Z03.1, Z08-Z09.9, Z12-Z12.9, Z80-Z80.9, Z85-Z85.9, Z86.0-Z86.03 |
| Neurological disorders | F00-F02.0, F02.2-F02.3, F02.8-F03.91, F06.2, G10-G10.0, G11-G13.8, G20-G21, G21.2-G24, G24.1-G25.0, G25.2-G25.3, G25.5, G25.8-G26.0, G30-G31.1, G31.8-G32.89, G35-G35.0, G36-G37.9, G40-G41.9, G43-G44.89, G50-G54.1, G54.5-G62, G62.2-G65.2, G70-G71.19, G71.3-G72, G72.1-G73.7, G80-G83.9, G89-G93.6, G93.8-G95.29, G95.8-G96, G96.1, G96.12-G96.9, G98-G99.8, M33-M33.99, M60-M60.19, M60.8-M60.9, M79.7, R25-R27.9, R29-R29.91, R41-R42.0, R56-R56.9, R90-R90.89, Z03.3, Z13.85, Z13.858, Z82.0, Z86.6-Z86.69 |
| Chronic respiratory disease | 189D86-D86.2, D86.9, G47.3-G47.39, J30-J35.9, J37-J39.9, J41-J42.4, J43-J46.0, J47-J47.9, J60-J68.9, J70.8-J70.9, J80-J80.9, J82, J84-J84.9, J90-J90.0, J91, J91.8-J93.12, J93.8-J94.9, J96-J96.92, J98-J99.8, R05.0-R06.9, R09-R09.89, R84-R84.9, R91-R91.8, Z82.5 |
| Diabetes and kidney disease | D63.1, E08-E08.9, E10-E14.9, I12-I13.9, N00-N08.8, N15.0, N17-N19, Q60-Q63.2, Q63.8-Q63.9, Q64.2-Q64.9, R73-R73.9, Z13.1, Z49-Z49.32, Z52.4, Z83.3, Z99.2 |

**S3 Table.**
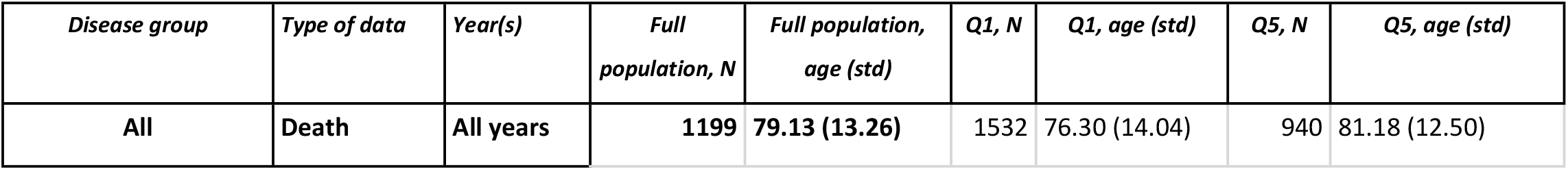
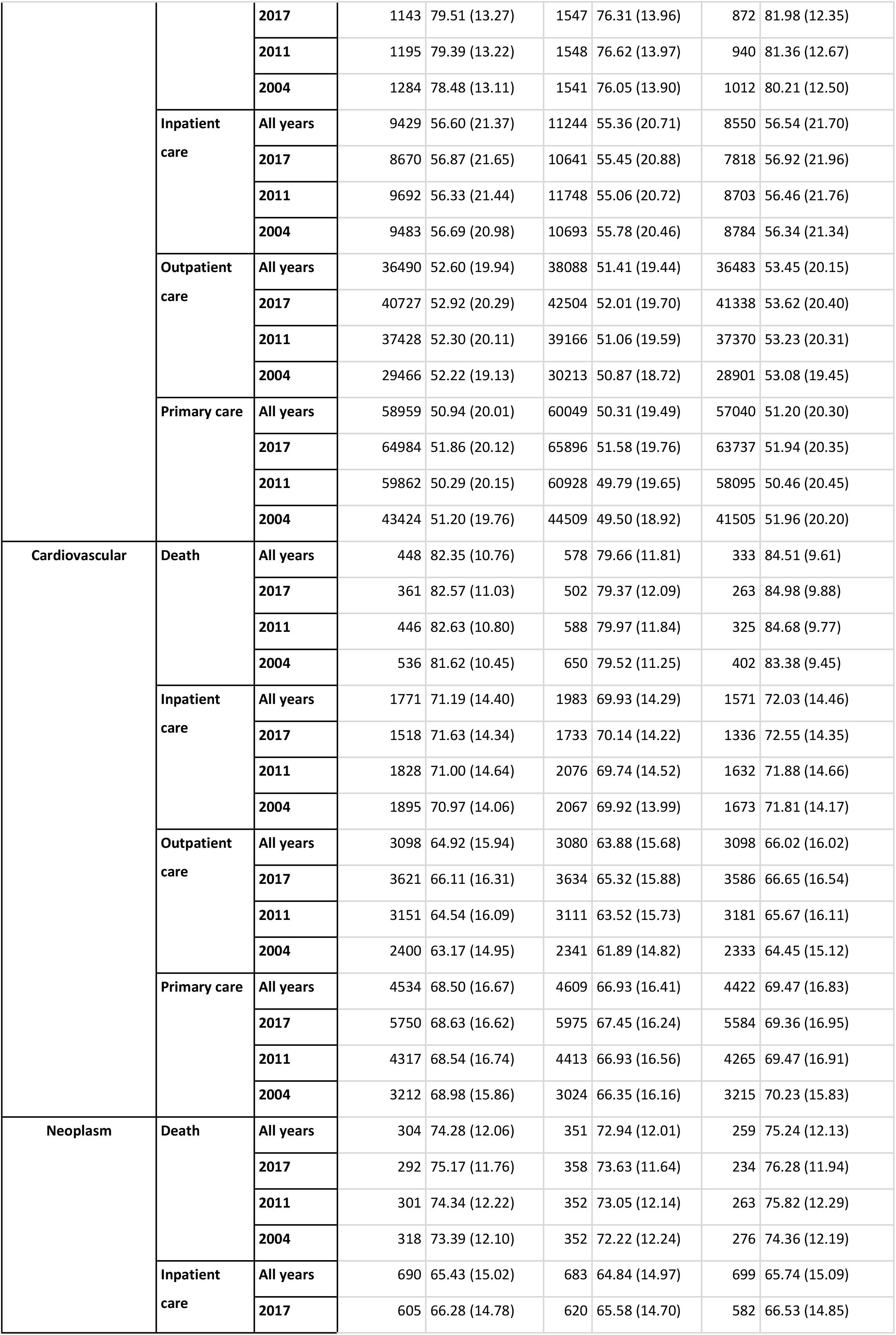
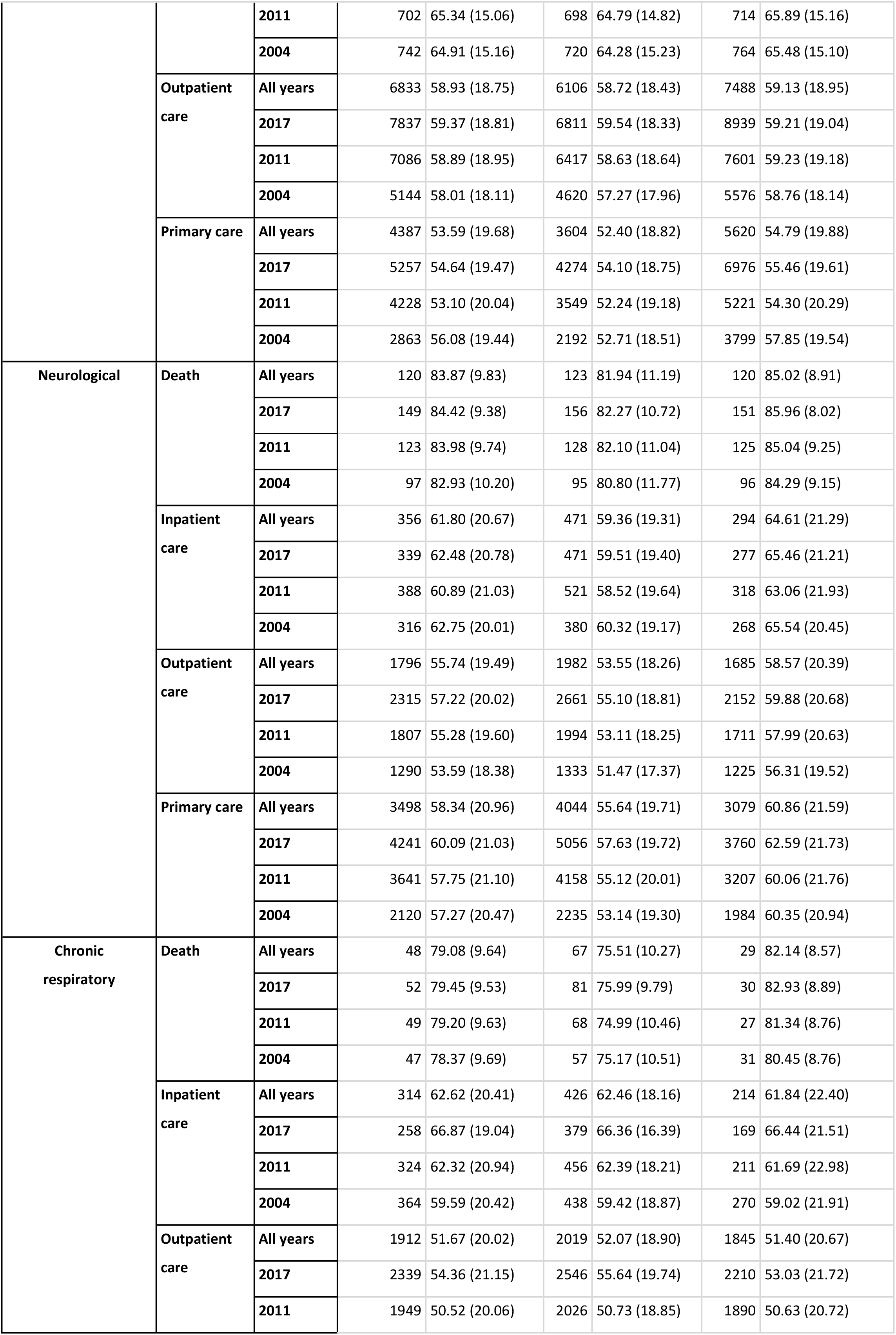
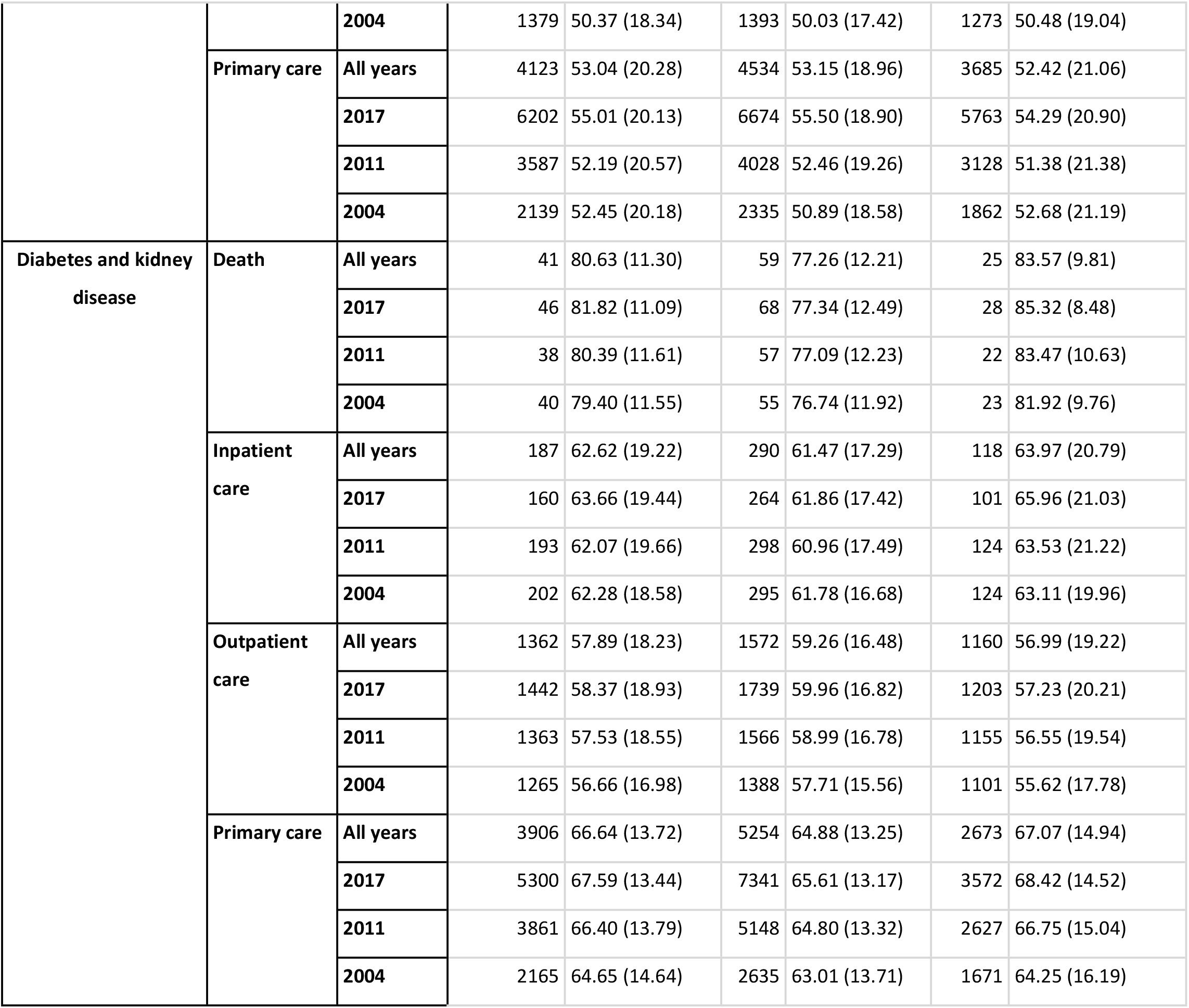
Rates (per 100 000) of deaths and diagnosed unique individuals by disease group, type of data, year, for the total sample and for each income group (Q1 and Q5). (Since socioeconomic data were available only for fully survived calendar years, dates of death are projected to the previous year, hence rendering systematic underestimation of true age of death).

**S4 Table.** Adjusted OR (Q1/Q5) in 2017 and yearly change of log transformed OR.

| Disease category | Type of data | OR (95% CI), 2004 | OR (95% CI ), 2017 | Yearly change in OR (95% CI) |
| --- | --- | --- | --- | --- |
| All | Death | 1.504 (1.471,1.538) | 1.776 (1.737,1.816) | 0.013 (0.012,0.015) |
|  | Inpatient care | 1.231 (1.221,1.241) | 1.436 (1.425,1.448) | 0.012 (0.012,0.013) |
|  | Outpatient care | 1.038 (1.033,1.044) | 1.042 (1.037,1.047) | -0.006 (-0.007,-0.006) |
|  | PC_Primary careu | 1.113 (1.106,1.120) | 1.071 (1.065,1.077) | -0.003 (-0.003,-0.002) |
| Cardiovascular | Death | 1.570 (1.517,1.625) | 1.820 (1.751,1.892) | 0.012 (0.010,0.015) |
|  | Inpatient care | 1.213 (1.191,1.235) | 1.288 (1.264,1.313) | 0.007 (0.005,0.008) |
|  | Outpatient care | 1.001 (0.986,1.017) | 1.051 (1.038,1.064) | 0.001 (0.000,0.002) |
|  | Primary care | 0.974 (0.957,0.992) | 1.073 (1.061,1.084) | 0.008 (0.007,0.009) |
| Neoplasm | Death | 1.261 (1.208,1.316) | 1.536 (1.471,1.603) | 0.017 (0.014,0.020) |
|  | Inpatient care | 0.959 (0.932,0.986) | 1.095 (1.063,1.128) | 0.005 (0.003,0.007) |
|  | Outpatient care | 0.840 (0.831,0.850) | 0.783 (0.777,0.790) | -0.013 (-0.013,-0.012) |
|  | Primary care | 0.545 (0.534,0.555) | 0.576 (0.570,0.583) | -0.003 (-0.004,-0.002) |
| Neurological | Death | 0.974 (0.902,1.052) | 1.001 (0.945,1.061) | -0.001 (-0.006,0.004) |
|  | Inpatient care | 1.364 (1.307,1.423) | 1.687 (1.622,1.753) | 0.016 (0.012,0.019) |
|  | Outpatient care | 1.076 (1.054,1.099) | 1.282 (1.263,1.302) | 0.012 (0.011,0.014) |
|  | Primary care | 1.097 (1.073,1.121) | 1.286 (1.271,1.302) | 0.016 (0.015,0.018) |
| Chronic respiratory | Death | 1.670 (1.485,1.877) | 2.438 (2.190,2.715) | 0.024 (0.016,0.032) |
|  | Inpatient care | 1.537 (1.475,1.601) | 2.129 (2.032,2.232) | 0.027 (0.024,0.030) |
|  | Outpatient care | 1.055 (1.033,1.077) | 1.098 (1.081,1.115) | -0.006 (-0.007,-0.005) |
|  | Primary care | 1.178 (1.153,1.203) | 1.108 (1.096,1.119) | -0.011 (-0.012,-0.010) |
| Diabetes and kidney disease | Death | 2.262 (1.984,2.580) | 2.170 (1.940,2.426) | 0.000 (-0.009,0.009) |
|  | Inpatient care | 2.223 (2.100,2.353) | 2.439 (2.298,2.589) | 0.010 (0.006,0.014) |
|  | Outpatient care | 1.231 (1.204,1.258) | 1.465 (1.437,1.494) | 0.010 (0.008,0.011) |
|  | Primary care | 1.512 (1.480,1.545) | 1.954 (1.932,1.977) | 0.020 (0.019,0.021) |

**S5 Table.**
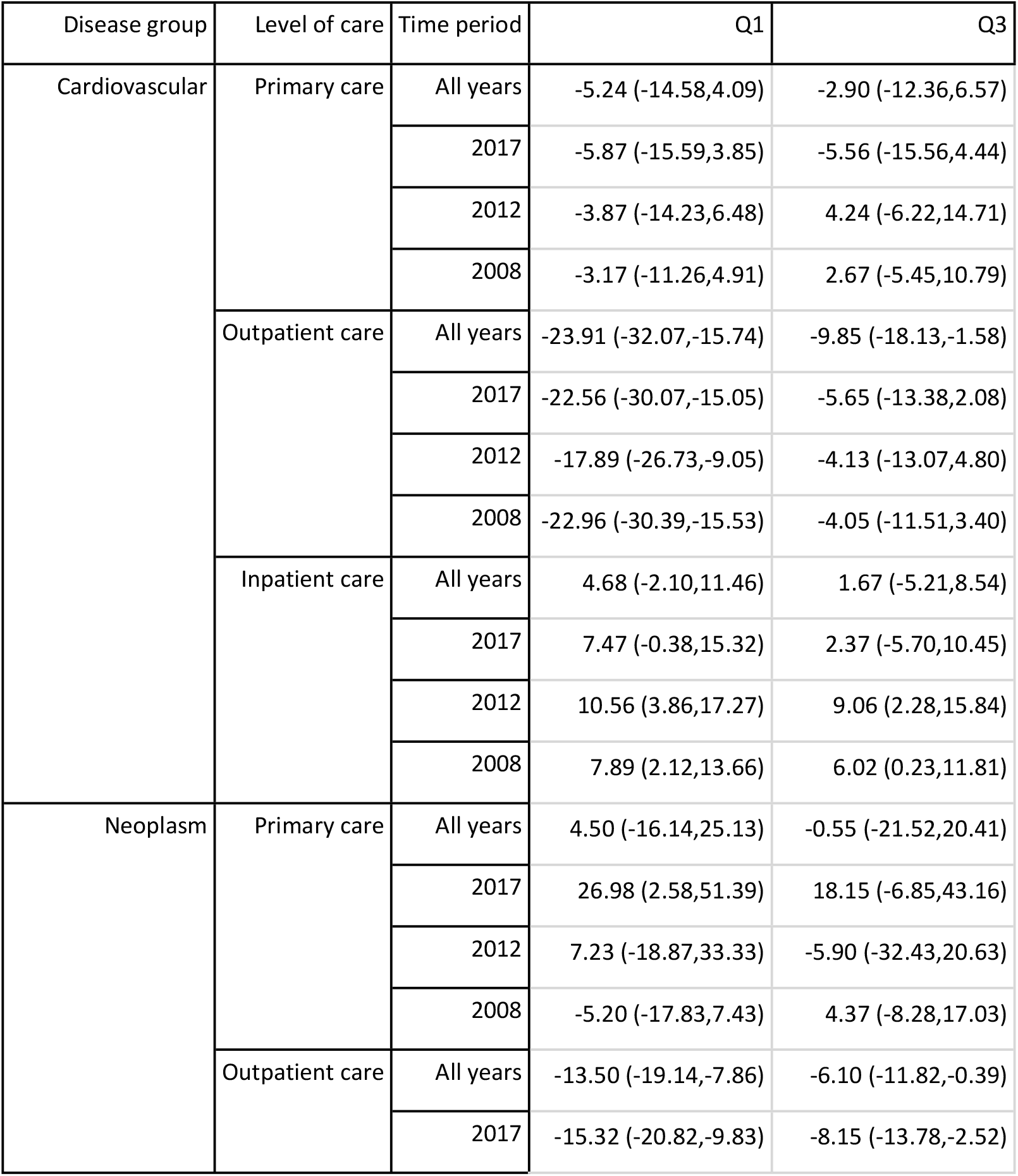
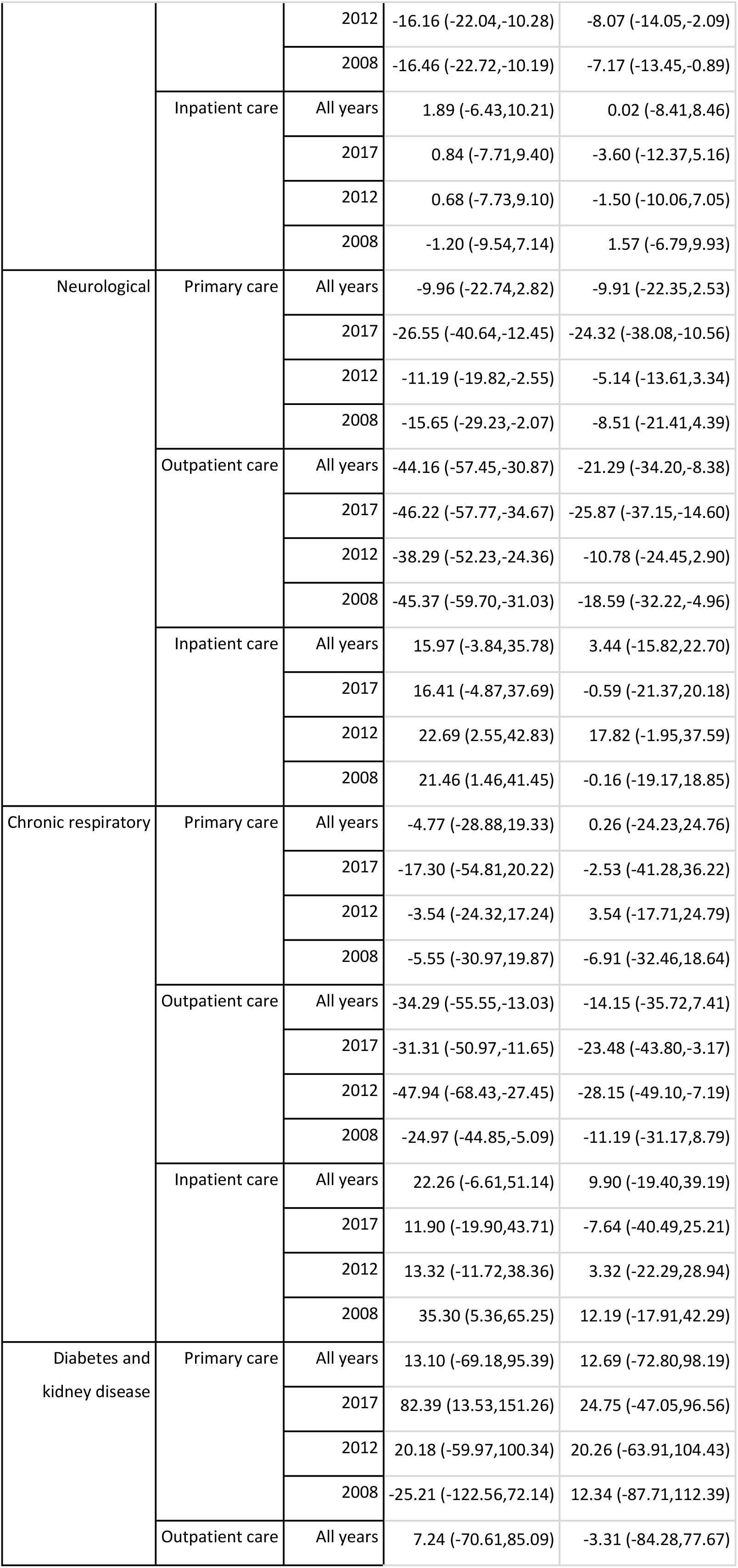
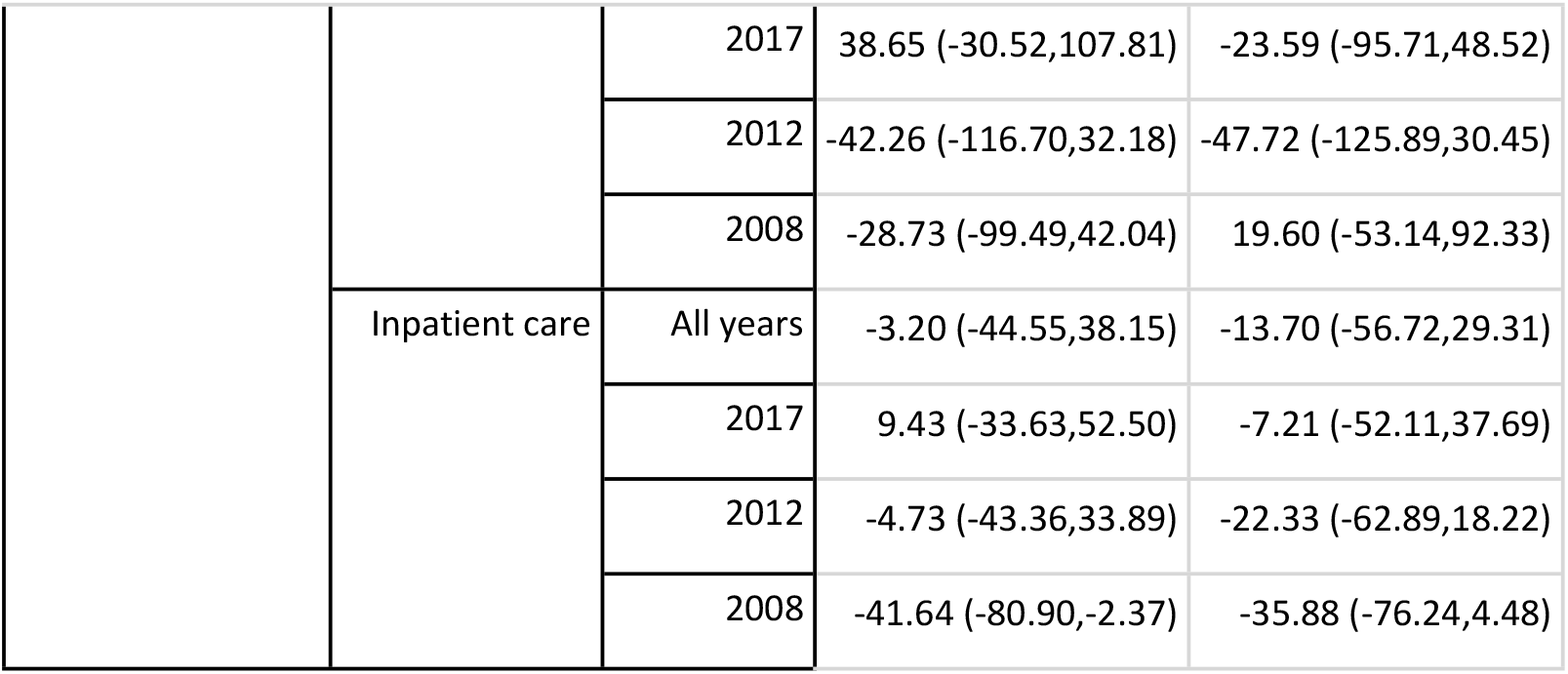
Health care visits within 5 years prior to death, by income. Differences in the number of health care encounters related to the cause of death. Differences are expressed as percentage change of the number of encounters in the highest income quintile.

| Disease group | Level of care | Time period | Q1 | Q3 |
| --- | --- | --- | --- | --- |
| Cardiovascular | Primary care | All years | -5.24 (-14.58,4.09) | -2.90 (-12.36,6.57) |
|  |  | 2017 | -5.87 (-15.59,3.85) | -5.56 (-15.56,4.44) |
|  |  | 2012 | -3.87 (-14.23,6.48) | 4.24 (-6.22,14.71) |
|  |  | 2008 | -3.17 (-11.26,4.91) | 2.67 (-5.45,10.79) |
|  | Outpatient care | All years | -23.91 (-32.07,-15.74) | -9.85 (-18.13,-1.58) |
|  |  | 2017 | -22.56 (-30.07,-15.05) | -5.65 (-13.38,2.08) |
|  |  | 2012 | -17.89 (-26.73,-9.05) | -4.13 (-13.07,4.80) |
|  |  | 2008 | -22.96 (-30.39,-15.53) | -4.05 (-11.51,3.40) |
|  | Inpatient care | All years | 4.68 (-2.10,11.46) | 1.67 (-5.21,8.54) |
|  |  | 2017 | 7.47 (-0.38,15.32) | 2.37 (-5.70,10.45) |
|  |  | 2012 | 10.56 (3.86,17.27) | 9.06 (2.28,15.84) |
|  |  | 2008 | 7.89 (2.12,13.66) | 6.02 (0.23,11.81) |
| Neoplasm | Primary care | All years | 4.50 (-16.14,25.13) | -0.55 (-21.52,20.41) |
|  |  | 2017 | 26.98 (2.58,51.39) | 18.15 (-6.85,43.16) |
|  |  | 2012 | 7.23 (-18.87,33.33) | -5.90 (-32.43,20.63) |
|  |  | 2008 | -5.20 (-17.83,7.43) | 4.37 (-8.28,17.03) |
|  | Outpatient care | All years | -13.50 (-19.14,-7.86) | -6.10 (-11.82,-0.39) |
|  |  | 2017 | -15.32 (-20.82,-9.83) | -8.15 (-13.78,-2.52) |
|  |  | 2012 | -16.16 (-22.04,-10.28) | -8.07 (-14.05,-2.09) |
|  |  | 2008 | -16.46 (-22.72,-10.19) | -7.17 (-13.45,-0.89) |
|  | Inpatient care | All years | 1.89 (-6.43,10.21) | 0.02 (-8.41,8.46) |
|  |  | 2017 | 0.84 (-7.71,9.40) | -3.60 (-12.37,5.16) |
|  |  | 2012 | 0.68 (-7.73,9.10) | -1.50 (-10.06,7.05) |
|  |  | 2008 | -1.20 (-9.54,7.14) | 1.57 (-6.79,9.93) |
| Neurological | Primary care | All years | -9.96 (-22.74,2.82) | -9.91 (-22.35,2.53) |
|  |  | 2017 | -26.55 (-40.64,-12.45) | -24.32 (-38.08,-10.56) |
|  |  | 2012 | -11.19 (-19.82,-2.55) | -5.14 (-13.61,3.34) |
|  |  | 2008 | -15.65 (-29.23,-2.07) | -8.51 (-21.41,4.39) |
|  | Outpatient care | All years | -44.16 (-57.45,-30.87) | -21.29 (-34.20,-8.38) |
|  |  | 2017 | -46.22 (-57.77,-34.67) | -25.87 (-37.15,-14.60) |
|  |  | 2012 | -38.29 (-52.23,-24.36) | -10.78 (-24.45,2.90) |
|  |  | 2008 | -45.37 (-59.70,-31.03) | -18.59 (-32.22,-4.96) |
|  | Inpatient care | All years | 15.97 (-3.84,35.78) | 3.44 (-15.82,22.70) |
|  |  | 2017 | 16.41 (-4.87,37.69) | -0.59 (-21.37,20.18) |
|  |  | 2012 | 22.69 (2.55,42.83) | 17.82 (-1.95,37.59) |
|  |  | 2008 | 21.46 (1.46,41.45) | -0.16 (-19.17,18.85) |
| Chronic respiratory | Primary care | All years | -4.77 (-28.88,19.33) | 0.26 (-24.23,24.76) |
|  |  | 2017 | -17.30 (-54.81,20.22) | -2.53 (-41.28,36.22) |
|  |  | 2012 | -3.54 (-24.32,17.24) | 3.54 (-17.71,24.79) |
|  |  | 2008 | -5.55 (-30.97,19.87) | -6.91 (-32.46,18.64) |
|  | Outpatient care | All years | -34.29 (-55.55,-13.03) | -14.15 (-35.72,7.41) |
|  |  | 2017 | -31.31 (-50.97,-11.65) | -23.48 (-43.80,-3.17) |
|  |  | 2012 | -47.94 (-68.43,-27.45) | -28.15 (-49.10,-7.19) |
|  |  | 2008 | -24.97 (-44.85,-5.09) | -11.19 (-31.17,8.79) |
|  | Inpatient care | All years | 22.26 (-6.61,51.14) | 9.90 (-19.40,39.19) |
|  |  | 2017 | 11.90 (-19.90,43.71) | -7.64 (-40.49,25.21) |
|  |  | 2012 | 13.32 (-11.72,38.36) | 3.32 (-22.29,28.94) |
|  |  | 2008 | 35.30 (5.36,65.25) | 12.19 (-17.91,42.29) |
| Diabetes and kidney disease | Primary care | All years | 13.10 (-69.18,95.39) | 12.69 (-72.80,98.19) |
|  |  | 2017 | 82.39 (13.53,151.26) | 24.75 (-47.05,96.56) |
|  |  | 2012 | 20.18 (-59.97,100.34) | 20.26 (-63.91,104.43) |
|  |  | 2008 | -25.21 (-122.56,72.14) | 12.34 (-87.71,112.39) |
|  | Outpatient care | All years | 7.24 (-70.61,85.09) | -3.31 (-84.28,77.67) |
|  |  | 2017 | 38.65 (-30.52,107.81) | -23.59 (-95.71,48.52) |
|  |  | 2012 | -42.26 (-116.70,32.18) | -47.72 (-125.89,30.45) |
|  |  | 2008 | -28.73 (-99.49,42.04) | 19.60 (-53.14,92.33) |
|  | Inpatient care | All years | -3.20 (-44.55,38.15) | -13.70 (-56.72,29.31) |
|  |  | 2017 | 9.43 (-33.63,52.50) | -7.21 (-52.11,37.69) |
|  |  | 2012 | -4.73 (-43.36,33.89) | -22.33 (-62.89,18.22) |
|  |  | 2008 | -41.64 (-80.90,-2.37) | -35.88 (-76.24,4.48) |

## Notes

### Competing Interest Statement

The authors have declared no competing interest.

### Funding Statement

Initials of the authors who received each award: E.A Grant numbers awarded to each author: DNR: 2021-00176 The full name of each funder: Swedish Research Council for Health, Working Life and Welfare URL of each funder website:https://www.vr.se/english.html The funders had no role in study design, data collection and analysis, decision to publish, or preparation of the manuscript.

### Author Declarations

Ethical standards: The authors assert that all procedures contributing to this work comply with the ethical standards of the relevant national and institutional committees on human experimentation and with the Helsinki Declaration of 1975, as revised in 2008. The registration number for the ethical applications and approvals by Regional Ethical Review Board, Stockholm, Sweden are: DNR 2018/1339-31/5, 2018/2292-32, 2019-02185, 2021-00657 and 2022-03111-02.

